# Assessment of COVID-19 Pandemic in Nepal: A Lockdown Scenario Analysis

**DOI:** 10.1101/2020.09.03.20187807

**Authors:** Kusum Sharma, Amrit Banstola, Rishi Ram Parajuli

**Affiliations:** Science Hub, Kathmandu, Nepal; Department of Research and Training, Public Health Perspective Nepal, Pokhara, Nepal; Faculty of Health and Applied Sciences, University of the West of England, Bristol, UK; Department of Civil Engineering, University of Bristol, UK

**Keywords:** COVID-19, coronavirus, public health, Nepal, spatial distribution, lockdown, impacts, challenges

## Abstract

The Government of Nepal issued a nationwide lockdown from 24 March to 21 July 2020, prohibiting domestic and international travels, closure of the border and non-essential services. There were only two confirmed cases from 610 Reverse Transcription Polymerase Chain Reaction (RT-PCR) tests and no fatalities when the government introduced nationwide lockdown. This study aimed to explore the overall scenario of COVID-19 including spatial distribution of cases; government efforts, and impact on public health, socio-economy, and education during the lockdown in Nepal. We collated and analysed data using official figures from the Nepalese Ministry of Health and Population. Nepal had performed 7,791 RT-PCR tests for COVID-19, the highest number of tests during the lockdown. It has recorded its highest daily rise in coronavirus infections with a total of 740 new cases from the total of 4,483 RT-PCR tests performed on a single day. Nepal had reported a total of 17,994 positive cases and 40 deaths at the end of lockdown. The spatial distribution clearly shows that the cases were rapidly spreading from the southern part of the country where most points of entry and exit from India are located. To contain the spread of the virus, the government has also initiated various preventive measures and strategies during the lockdown. The Government of Nepal needs to allocate more resources, increase its capacity to test and trace, establish dedicated isolation and quarantine facility and impose local restrictions such as a local lockdown based on risk assessment rather than a nationwide lockdown.

## 1 INTRODUCTION

Coronavirus disease (COVID-19) outbreak originating from Wuhan, China in late 2019 has spread worldwide claiming more than 2.5 million lives all over the world as of 01 March 2021 (1). On 11 March 2020, the World Health Organization (WHO) declared it as a pandemic (1). Since the outbreak of the disease WHO through its guidelines has prioritised the actions for responding to the virus; urged the government to maintain health facilities, raise public awareness, and stock up on medical supplies (2).

Several modelling studies have been conducted during the early phases of the outbreak to predict the epidemic and effectiveness of multiple population-wide strategies, including lockdown, social distancing, quarantine, testing and contact tracing, and media-related awareness among others to mitigate the spread of COVID-19 (3-9). The strict lockdown was enforced to limit the spread of COVID-19 in countries such as Italy, Spain, France, the UK after the steady rise in cases whereas Nepal introduced lockdown during the early phase of the pandemic (10). Lockdown is the blanket approach that buys time to prepare the healthcare system (active case finding through testing and tracing, case management e.g. quarantine, isolation and treatment, and availability of protective equipment) to confine the virus and its spread. The Government of Nepal issued a nationwide lockdown from 24 March to 21 July 2020, prohibiting domestic and international travels, closure of border and non-essential services in the first stage, which was later eased on 11 June 2020.

The basic reproduction number (R0) which measures the potential transmission of an infectious disease is a fundamental metric to determine if an outbreak is expected to continue. In general, the disease is expected to spread and become epidemic if R0 is more than one and to decline and ultimately end if R0 is less than one. The R0 value of the COVID-19 outbreak was not available for Nepal when the government was preparing for the lockdown. However, in neighbouring countries, the estimated R0 value of coronavirus was above 2.0 in India (4) and between 2.2 to 3.5 in China (11), indicating a potential to cause an outbreak. There were only two confirmed cases from 610 Reverse Transcription Polymerase Chain Reaction (RT-PCR) tests and no fatalities before lockdown (12). The indexed case was found on 23 January 2020 in Kathmandu on a person who had travelled from Wuhan, China (13). The second case was confirmed two months later on 23 March who had travelled to Nepal from France via Qatar (14).

This study aimed to assess the overall scenario of COVID-19 during the lockdown (positive cases, RT-PCR test performed, recoveries, total active and deaths cases including case fatality ratio) including spatial distribution of the cases, government measures to manage the pandemic; and impacts on public health, socio-economy, and education. Finally, we offer helpful suggestions to address the challenges brought by the impact of COVID-19.

## 2 METHODS

### 2.1 Study Design

This was a descriptive study to assess the multiple scenarios of the COVID-19 pandemic in Nepal during the lockdown period. Statistical and spatial presentations of data, government efforts, impacts on public health, socio-economy, and education were discussed to summarize the situation of Nepal during the lockdown period. The main focus of the study was for the lockdown period, however, data was updated and discussed to reflect the post lockdown scenario.

### 2.2 Study Area

Nepal is a lower-middle-income country in South Asia between India (in the south, east, and west) and China (in the north). It has a population of around 30 million. The new constitution promulgated on 20 September 2015 made Nepal a federal democratic republic and is now divided into seven provinces, 77 districts, and 753 local units (municipalities and rural municipalities). It is divided into three physiographic regions: Mountain region (Great Himalayan Range in the northern part), Hilly region, and Terai region (low land region at the Indian border in the southern part).

### 2.3 Data Collection

We analysed results from the COVID-19 situation reports prepared by the Nepalese Ministry of Health and Population (MOHP). For this study, we downloaded the COVID-19 situation reports from the openly available online resources on the web portal (covid19.mohp.gov.np). We collated the data for the lockdown period (from 24 March to 21 July 2020). The MOHP has defined a confirmed COVID-19 case as any person with laboratory confirmation of COVID-19 infection, irrespective of clinical signs and symptoms, i.e. person who had RT-PCR tested positive for Severe Acute Respiratory Syndrome Coronavirus-2 (SARS-CoV-2). Municipal level data has been taken from the data prepared for *COVIRA*, a COVID-19 risk assessment tool (15).

### 2.4 Statistical Analysis

Daily positive cases reported in local administrative units (municipality or rural municipality) were analysed along with total daily data for the country. Data were analysed descriptively in a Microsoft Excel 2019 Version 16.0 (Microsoft Corporation, Redmond, Washington, USA). The number of COVID-19 cases, daily RT-PCR tests performed and the number of recoveries were used to assess the COVID-19 pandemic situation. The number of deaths and Case Fatality Rate (CFR) was presented by different age groups. CFR was calculated as the proportion of confirmed deaths among identified confirmed cases. Everyday data of COVID-19 positive cases were presented in local level units. The maps used to show the spatial distribution of COVID-19 cases in Nepal were created using the QGIS Version 3.14 (www.qgis.org).

## 3 RESULTS

Figure 1 shows the daily and weekly average of tests and positive cases during the lockdown period. On 29 June, Nepal had performed 7,791 tests for COVID-19, the highest number of tests during the lockdown. It has recorded its highest daily rise in coronavirus infections with a total of 740 new cases from the total of 4,483 test performed on a single day on 3 July. The number of daily cases was decreasing after its peak on 3 July but subsequently, the number of tests was also decreasing.

**Figure 1.**
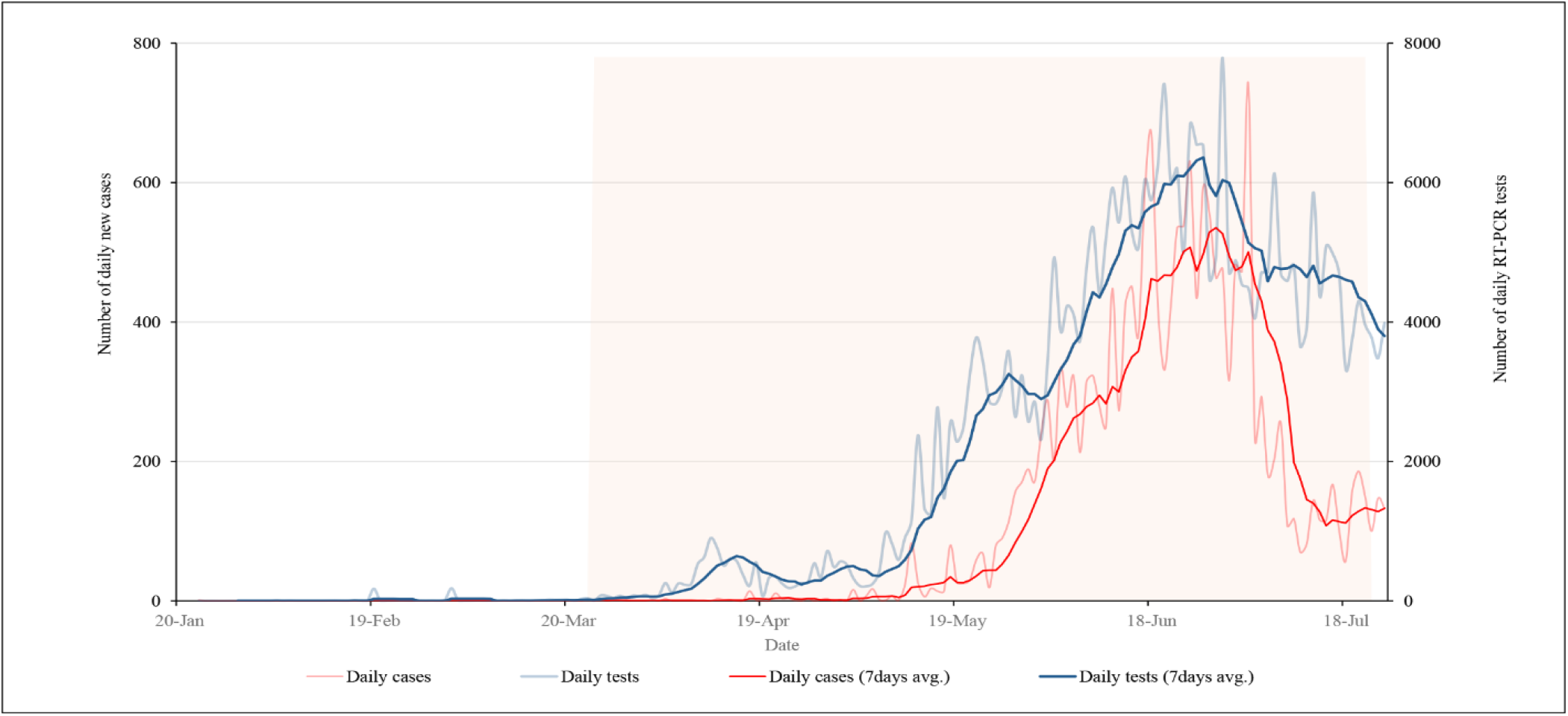
Daily new cases and RT-PCR tests along with 7 days average of cases and tests during the lockdown period.

Figure 2 shows COVID-19 cumulative cases, recoveries, and active cases during the lockdown. As per the figure, the number of people recovered from the coronavirus in Nepal increased during the lockdown. However, the number of active cases decreased. On 1 July there were 10,390 active cases in the country which was continuously declining till the end of the lockdown period. Lack of proper management of quarantine and isolation centres may have caused a significant increase in cases two-months after lockdown started as most of the cases were imported from India, and were later transmitted to the community. Data show that the bending of the curve had started before the government decided to lift the nationwide lockdown however the risks of further transmission in the community were prevailing.

**Figure 2.**
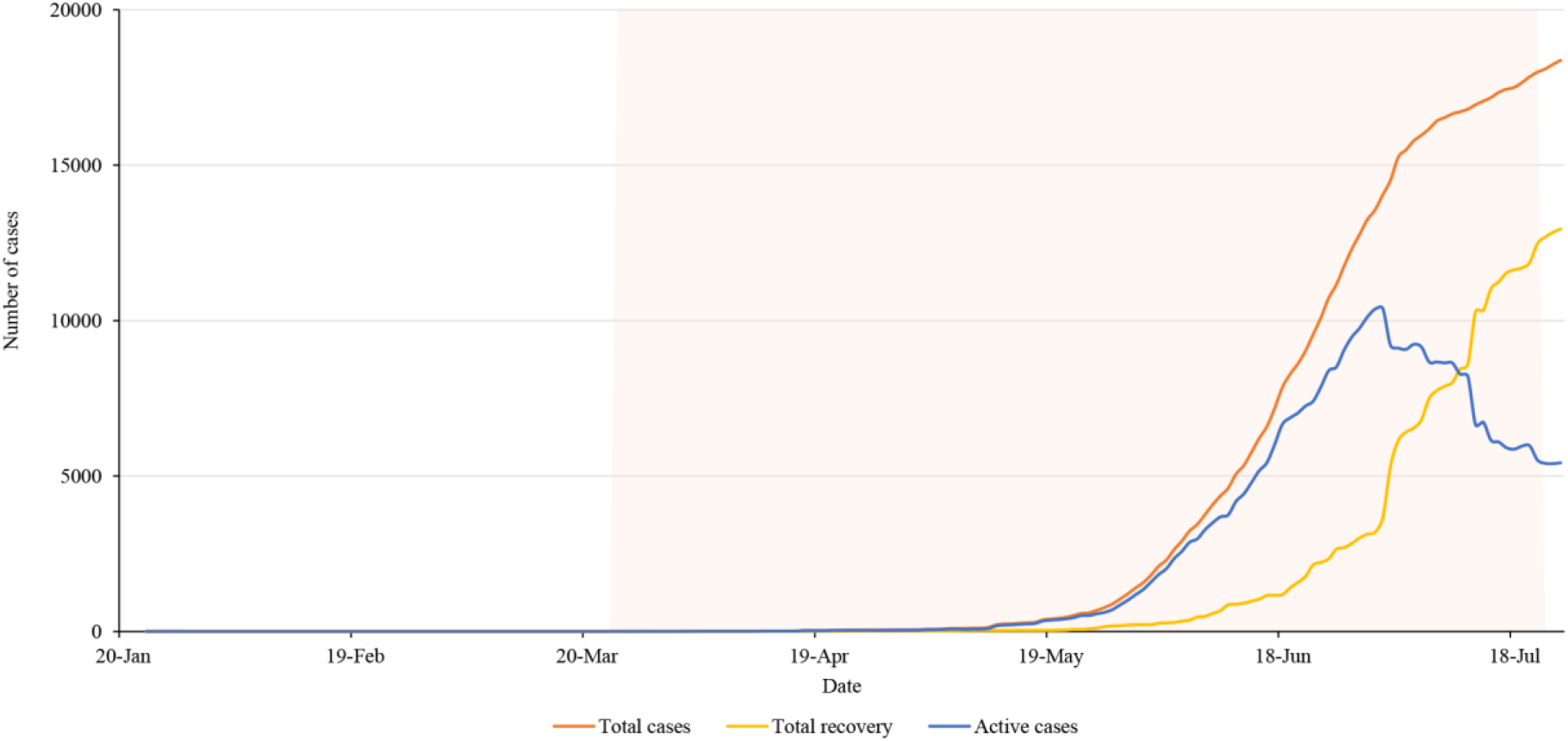
Total cases, recovery and active cases in Nepal during the lockdown period.

### 3.1 Death and Case Fatality Ratio

Nepal had reported a total of 17,994 positive cases and 40 deaths on the last date of nationwide lockdown. Figure 3 (a) and (b) shows the distribution of cases and a death toll respectively in age and gender groups. It clearly shows that the younger population of age group 21-30 years is more infected, and most importantly the fraction of female who got infected is relatively smaller. The main reason behind such distribution is most of the cases were imported by young immigrants who went abroad for work. The number of deaths and the corresponding overall death rate is very small (0.22%) however the relative death rate curve tells a different story (Figure 4).

**Figure 3(A).**
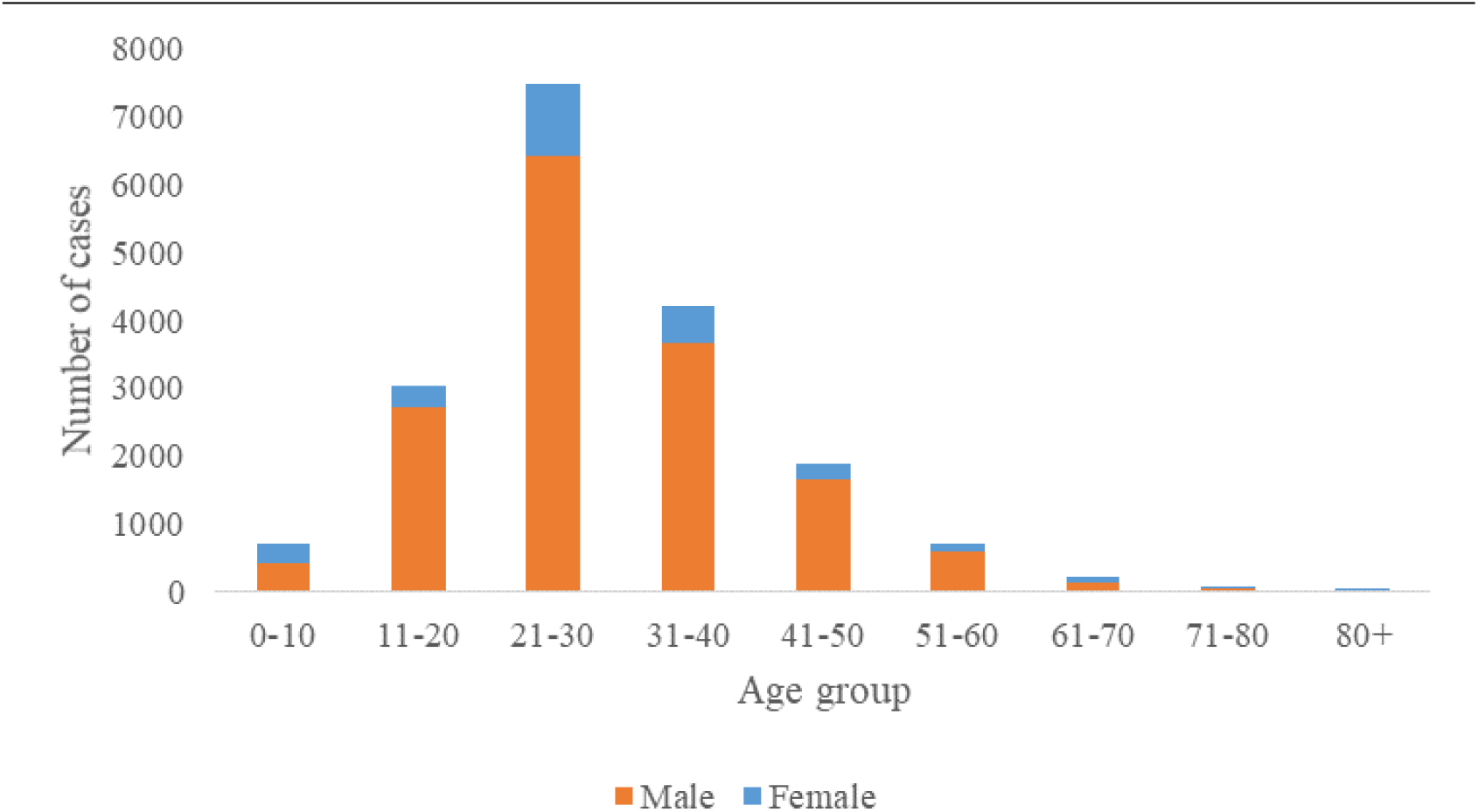
COVID-19 cases distribution among age and gender groups during the lockdown period.

**Figure 3 (B).**
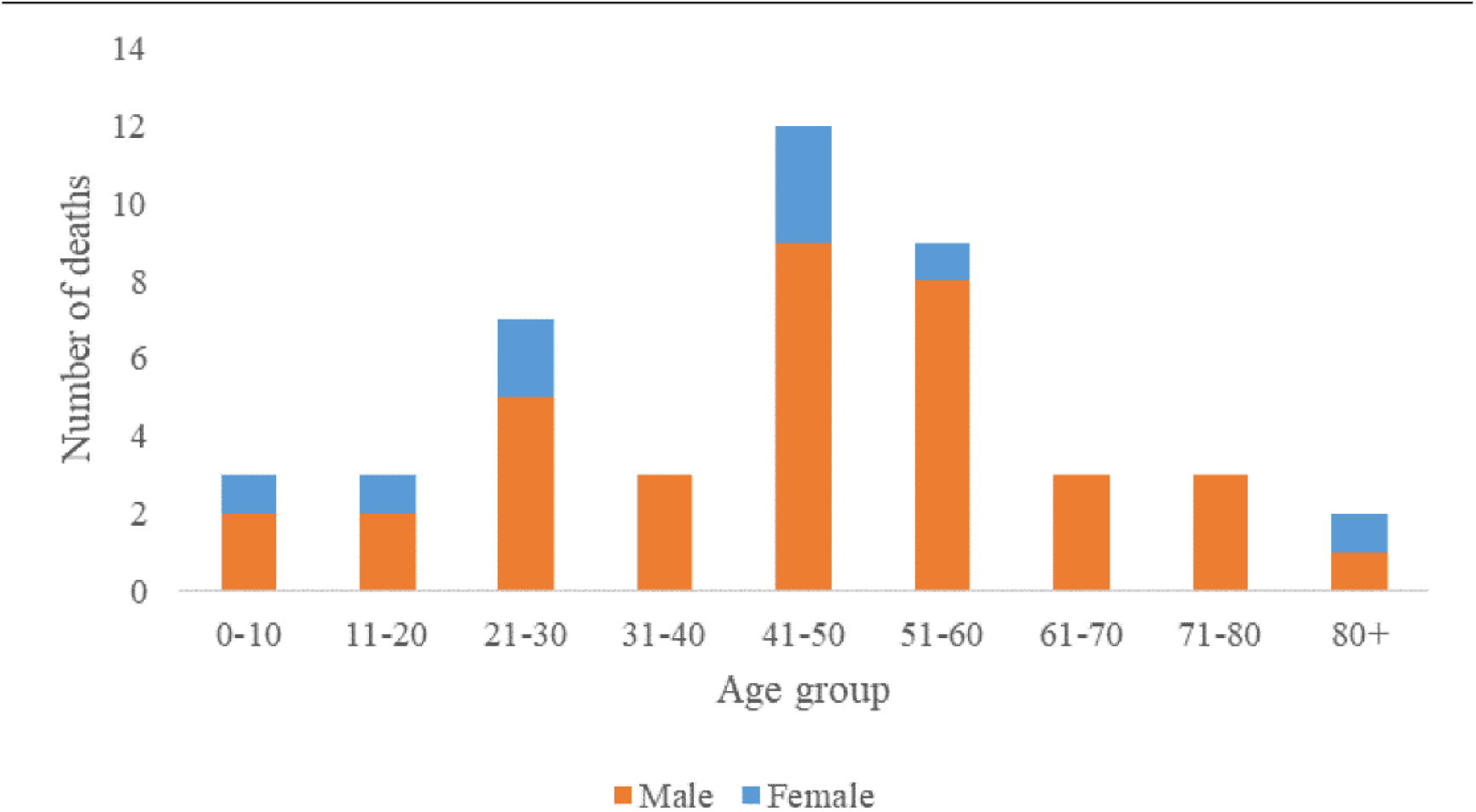
COVID-19 deaths distribution among age and gender groups during the lockdown period.

**Figure 4.**
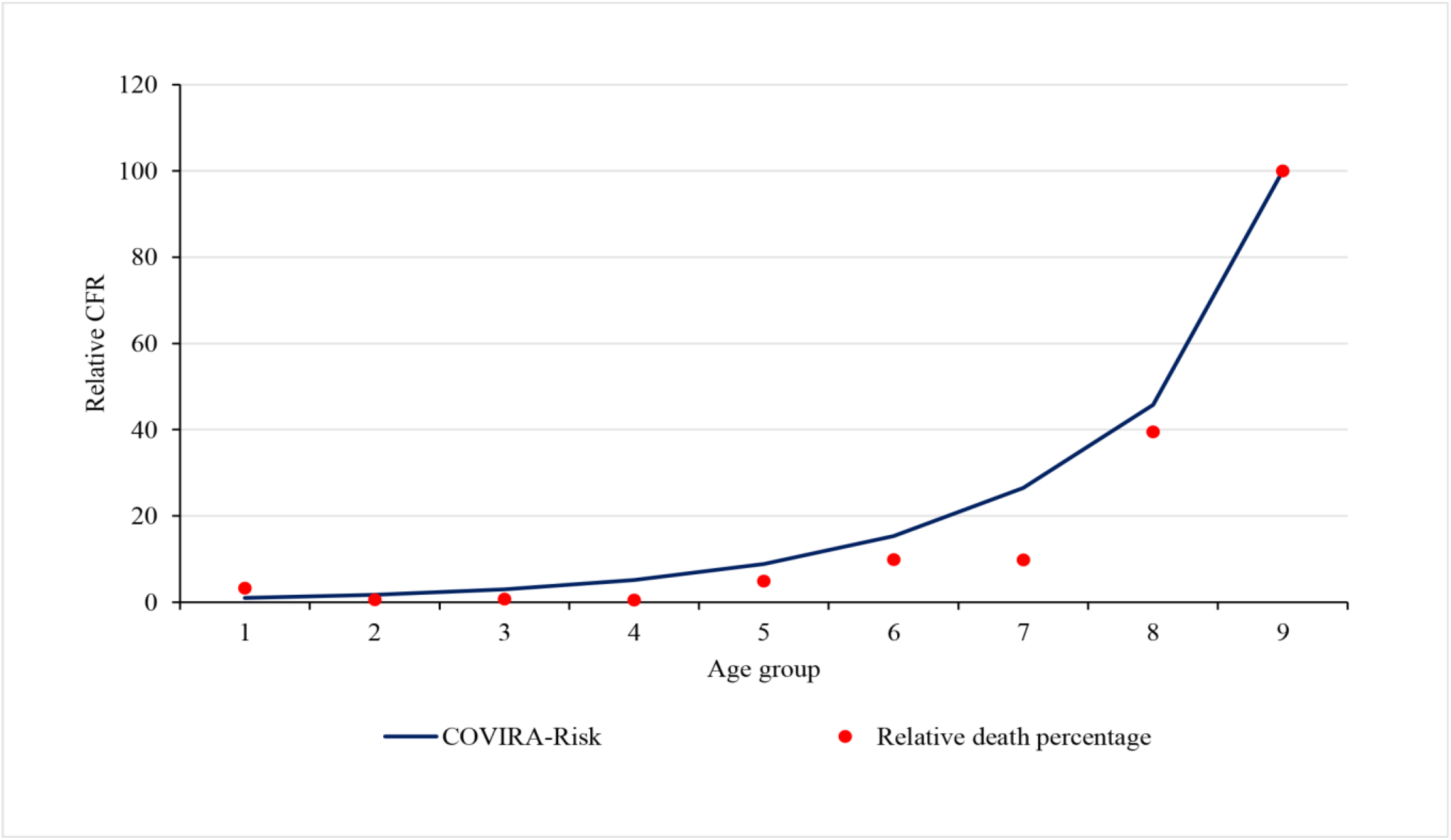
Relative case fatality ratio and comparison with the COVIRA risk model.

CFR in Nepal shows a similar trend to the *COVIRA* risk model (15) which is the risk model based on early-stage pandemic data. Figure 4 shows the comparison of the *COVIRA* risk model and Nepal data (relative percentage of CFR). The correlation between CFR in age groups and the *COVIRA* model is 0.986 which shows very good agreement of data. The trend of CFR by the age group shows an exponential relation with an r-square value of 0.714. Hence, one of the alarming points for Nepal is, if the infection spread in the community and the older people get infected, the death toll would rapidly increase.

### 3.2 Spatial Distribution

Figure 5 shows the spatial distribution of total cases on the first and the 15th days of the months over a lockdown period. These maps clearly show that the cases were rapidly spreading from the southern part of the country where most points of entry and exit from India are located.

**Figure 5.**
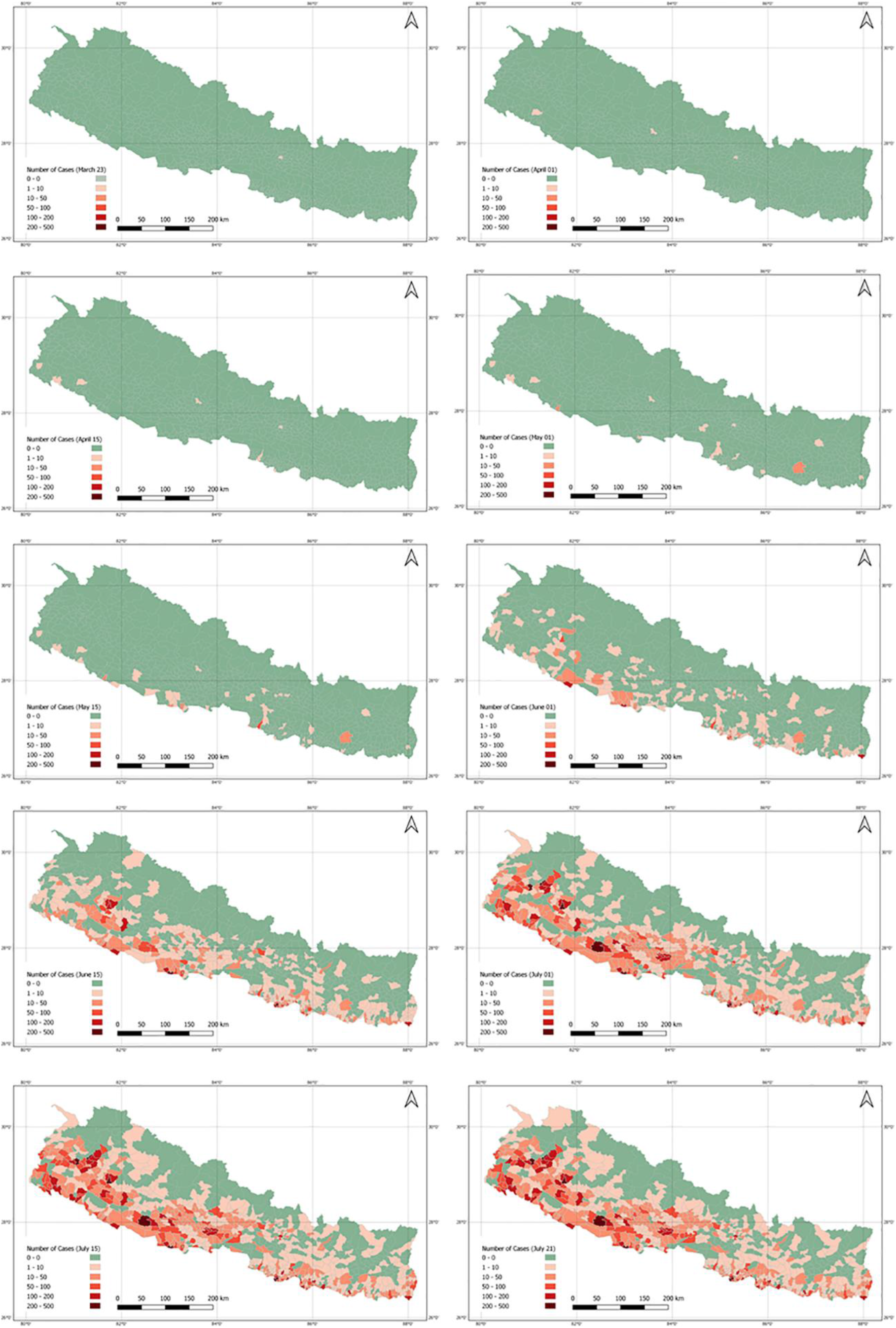
Total positive COVID-19 cases across the country by date during lockdown period.

The government lifted the lockdown when the curve of daily cases was flattening, however, the situation in the country was not totally under control. There was a high possibility of spreading the virus if an infected person was exposed to others until around two weeks of infection. Mapping the report of last active cases across the country was important to look at the prevailing risk zones. Figure 6 shows the number of days since the last case was reported on the day when the lockdown was lifted.

**Figure 6:**
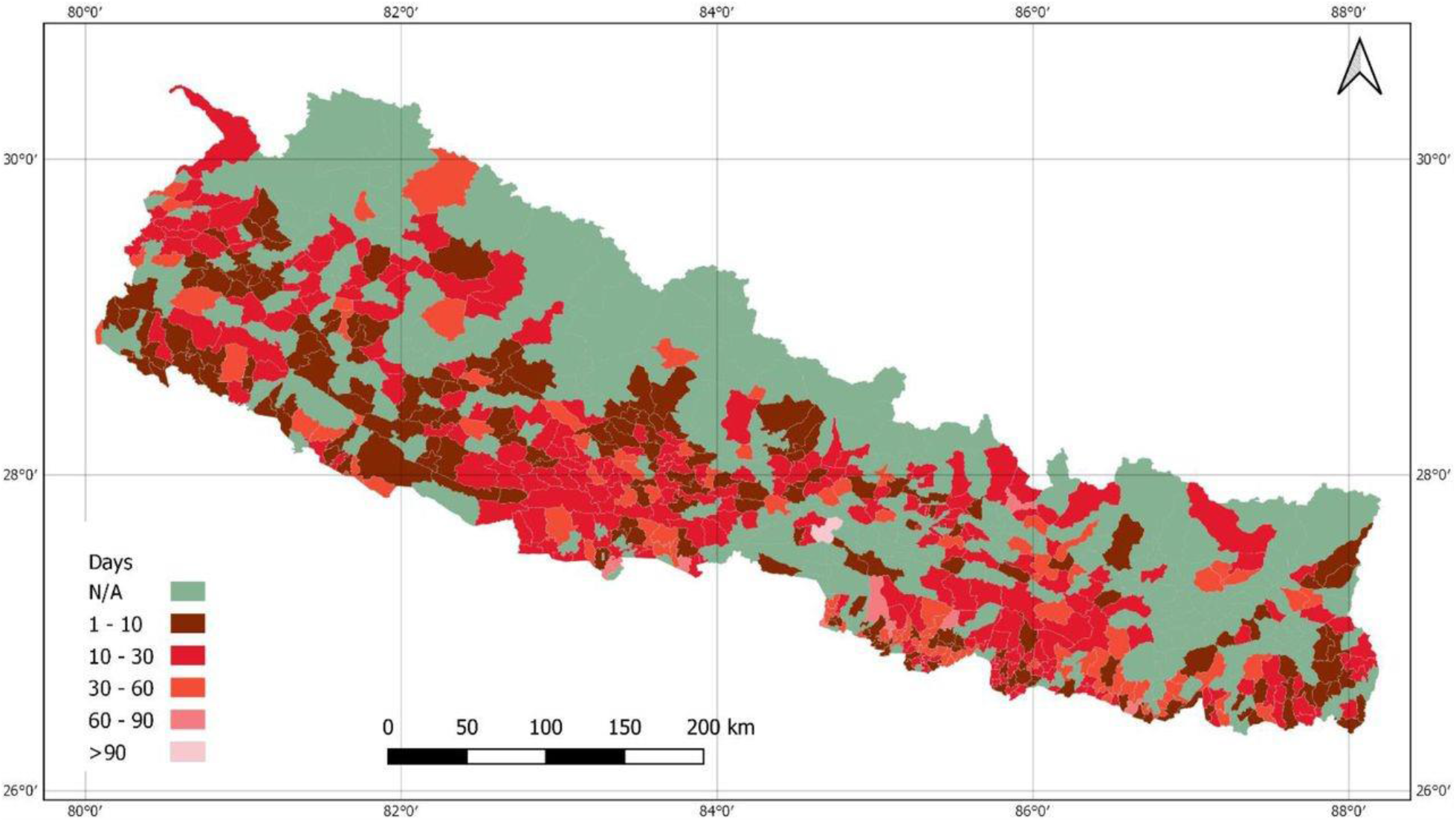
Number of days since the last case reported on the day of lockdown lifted.

### 3.3 Government Efforts on Managing the Pandemic During the Lockdown

The Government of Nepal has initiated various preventive measures and strategies during the lockdown as shown in the list below (8).

- Guidelines issued for the management and handling of quarantine.
- Dissemination of information, education and communication materials on social distancing, handwashing, proper use of masks and hand sanitizers, mass awareness via television, radio, social media and pamphlets.
- Launch of mobile application (*Hamro Swasthya*), the web portal (covid19.mohp.gov.np).
- Two toll-free call centres to provide counselling on COVID-19 prevention and treatment.
- Daily briefings by the MOHP to update about the current situation.
- Travel restriction, testing, and tracing.
- Health sector emergency response plan for COVID-19 pandemic which includes strategies to deal quarantine management, case investigation, contact tracing, community-level screening and testing, strengthening laboratory capacity etc.
- Protocol on the safe management of dead bodies.
- Guidelines issued for the management of isolation of COVID-19 cases.
- National testing guidelines for COVID-19.
- Public health standards to be followed by people and institutions during the COVID-19 pandemic and lockdown.
- Allowed private laboratories to perform RT-PCR test.
- Availability of RT-PCR tests in all seven provinces of Nepal.
- Health standards for isolation of COVID-19 cases.
- Training of trainers on case investigation and contact tracing.
- MOHP endorsed the standards for home quarantine.
- The number of hospitals for the management of COVID-19 (as of 21 July 2020):
  - Hospitals with COVID-19 clinics: 125
  - Level 1 COVID Hospitals (for management of positive cases with mild symptoms): 16
  - Level 2 COVID Hospitals (for management of positive cases with moderate or severe symptoms): 16
  - Level 3 COVID Hospitals (for management of COVID positive cases who needs multi-speciality services): 4
- Total laboratories established capable of doings RT-PCR: 28
- Total intensive care unit beds allocated for COVID-19 cases: 942 of total 2,600 ICU beds available in the country.
- Total ventilators allocated for COVID-19 cases: 496 of total 900 ventilators available in the country.

## 4 DISCUSSION

Nepal had performed 7,791 tests for COVID-19, the highest number of tests during the lockdown. It has recorded its highest daily rise in coronavirus infections with a total of 740 new cases from the total of 4,483 tests performed on a single day. The number of people recovered from the coronavirus in Nepal increased during the lockdown. Nepal had reported a total of 17,994 positive cases and 40 deaths on the last date of nationwide lockdown. The spatial distribution clearly shows that the cases were rapidly spreading from the southern part of the country where most points of entry and exit from India are located.

### 4.1 Socioeconomic Impact

Nationwide lockdown restricted the socioeconomic activities all over the country, where very few essential services were run throughout the period. Multidimensional impacts of lockdown have been found in society, many people lost their jobs, and businesses along with other health care were impacted. It disrupted the supply chain, shut many informal and small enterprises, and pushed more vulnerable people into poverty (17). The tourism industry hit hard in Nepal where it fell below 10%, resulting in more than 13,000 job loss of trekkers and guides (18). There are 1.6 to 2 million jobs at risk due to the COVID-19 crisis where 80.8% of total jobs in the country are informal (19). A household survey on the impact of COVID-19 on food security and vulnerability conducted by the World Food Programme, Nepal and the Ministry of Agriculture and Livestock Development showed that of the 4,416 households from across the country, only 42% had one month worth of food stocks (20).

### 4.2 Healthcare Impact

The lockdown has affected the health of individuals and disrupted healthcare services, particularly emergency and regular health services. During the lockdown, at the individual level, one of the most notable impacts was on psychological health. Quarantine, social isolation, and travel restrictions had negatively impacted the mental health of people who have COVID-19 and their families. A few preliminary studies have shown psychological issues such as stress, anxiety, depression, insomnia among the general population (21-23) as well as frontline health workers (24, 25). A study by Gupta and colleagues conducted among 150 health workers showed that 38 % of the healthcare workers on COVID-19 duty in Nepal suffered anxiety and/or depression (24). Another online survey conducted among 475 health workers showed that 41.9% of health workers had symptoms of anxiety, 37.5% had depression symptoms and 33.9% had symptoms of insomnia (25). Incidents of stigmatization and social discrimination of healthcare workers, people who have COVID-19 and their families were also reported in Nepal during the lockdown (26, 27).

These effects of lockdown on psychological health are in line with evidence from other countries. Studies from these countries show that lockdown, quarantine, and isolations have increased social isolation, frustration, loneliness, boredom, inadequate supplies, financial insecurity, and stigma which are associated with increased risk of depression, stress, anxiety, confusion, fear, emotional disturbance, insomnia, grief and irritability (28-30). A study conducted in India shows that the prevalence of depression and anxiety has increased by eight to tenfold among the adult population during lockdown (31). Women in general suffered more from lockdown reporting increased depression, anxiety, stress, and insomnia (32, 33).

Nepal Police record shows that during the lockdown, the number of suicide cases has increased. Within 74 days of lockdown, a total of 1,227 people committed suicide, which is more than 15 suicidal death per month compared to the previous year (34). Although reasons for what had caused suicide and suicidal thoughts are still unknown in Nepal, they could be linked to the uncertainty about the pandemic, self-isolation, financial burden, loss of family members, stigma as evident in previous disasters and epidemic (35-37). In addition to suicide, domestic violence, sexual abuse, and rape were being perpetrated during the lockdown in Nepal (38).

The government’s priority to combat COVID-19 and the lockdown adopted to contain its transmission put vulnerable populations such as pregnant women, children, the elderly and people with non-COVID diseases at risk by impacting their ability to access essential healthcare services. For example, pregnant women faced barriers to accessing regular antenatal care and delivery services (39) and patients with non-communicable diseases faced barriers to access long-term care and medicines (40) during the lockdown periods. Millions of children aged between six months and five years missed measles and rubella mass immunization, vitamin A, and deworming tablets because the Government of Nepal postponed these national-level campaigns (41). Limited ability to access such essential and routine health care services poses an urgent threat to the nation’s health and could reverse some of the achievements in reducing maternal, newborn, and child deaths.

### 4.3 Challenges

Future COVID-19 cases in Nepal will depend on the situation in India where the cases are increasing rapidly. Nepal shares an open border with India and there may be an increase in the number of Nepali workers returning from India who remain stranded in different parts of India due to the lockdowns in both countries. It is estimated that 600,000 migrant workers will return to Nepal within a few weeks of lifting the nationwide lockdown restrictions (42). This flow of migrant workers could increase the number of cases as the government has not been able to utilize the lockdown time efficiently to prepare and ready for responding to COVID-19. The challenge will be to test and trace these people for COVID-19. Before the lockdown, thousands of migrant Nepali workers returned to Nepal without proper screening from the Indian states of Maharashtra, Delhi and Gujarat, where the R0 value is more than one indicating an outbreak of COVID-19 in these states (4).

The testing capacity of the country has not increased due to a shortage of RT-PCR test kits, personal protective equipment, trained workers, and medical supplies. With limited testing capacity, it is challenging to monitor the transmission of the virus in Nepal because the suspected cases continue to transmit the virus while awaiting the COVID-19 test. Some COVID-19 cases remain asymptomatic, so it is difficult to predict the severity of the outbreak. There are only a few health facilities capable of treating and managing the cases with some degree of preparedness and readiness to provide the services (43, 44). If the number of cases becomes higher than the capacity of these health facilities to cope with the increased demand it would be more challenging to contain the virus.

Another challenge is to control other communicable and non-communicable diseases amidst the COVID-19 pandemic. Non-communicable diseases such as cancer, hypertension, cardiovascular disease, diabetes, chronic respiratory diseases, mental illness are already a major public health problem in Nepal and accounted for about 71% of the total annual deaths in 2019 (45). These diseases will exacerbate and the impact could be much higher than the COVID-19 if they are left behind in the fight against COVID-19.

### 4.4 Strengths and Limitations

The study strength included the use of openly available official figures from the MOHP web portal to provide an overall scenario of the COVID-19 pandemic during the lockdown. The findings would be applicable to compare the post-lockdown situation of the COVID-19 pandemic in Nepal and to guide more effective measures to contain the spread of the coronavirus. One of the limitations with openly available data was individual-level patient details were not accessible to perform detail epidemiological analysis. The number of people who had COVID-19 represents only a reflection of those who were tested rather than the actual figure. Also, manual gathering and submitting data from hospitals to the central government means can result in a delay and the loss of some information in reporting the number of deaths or cases so may not be a true reflection of the daily case counts. Further, the MoHP data exclude deaths from COVID-19 outside hospitals, such as those in the home. In the daily situation reports shared by the MoHP, there was inconsistency in the details provided. Some daily situation reports of the COVID-19 had information on gender while others did not contain such key information.

### 4.5 Post-lockdown scenario

This paper was mainly designed to present and discuss the lockdown scenario, however considering the publication date, data are updated and discussed for post-lockdown scenario. Figure 7 shows the RT-PCR tests including cases and Figure 8 shows the total cases, recoveries, and active cases as of 01 March 2021. The number of COVID-19 cases had increased until 21 October 2020 and after that the number of cases reported and number of tests conducted had declined (Figure 7). The two dips in Figure 7 i.e., on 27 October 2020 and 17 November 2020, were the festival days where less tests were performed and consequently less cases were identified.

**Figure 7:**
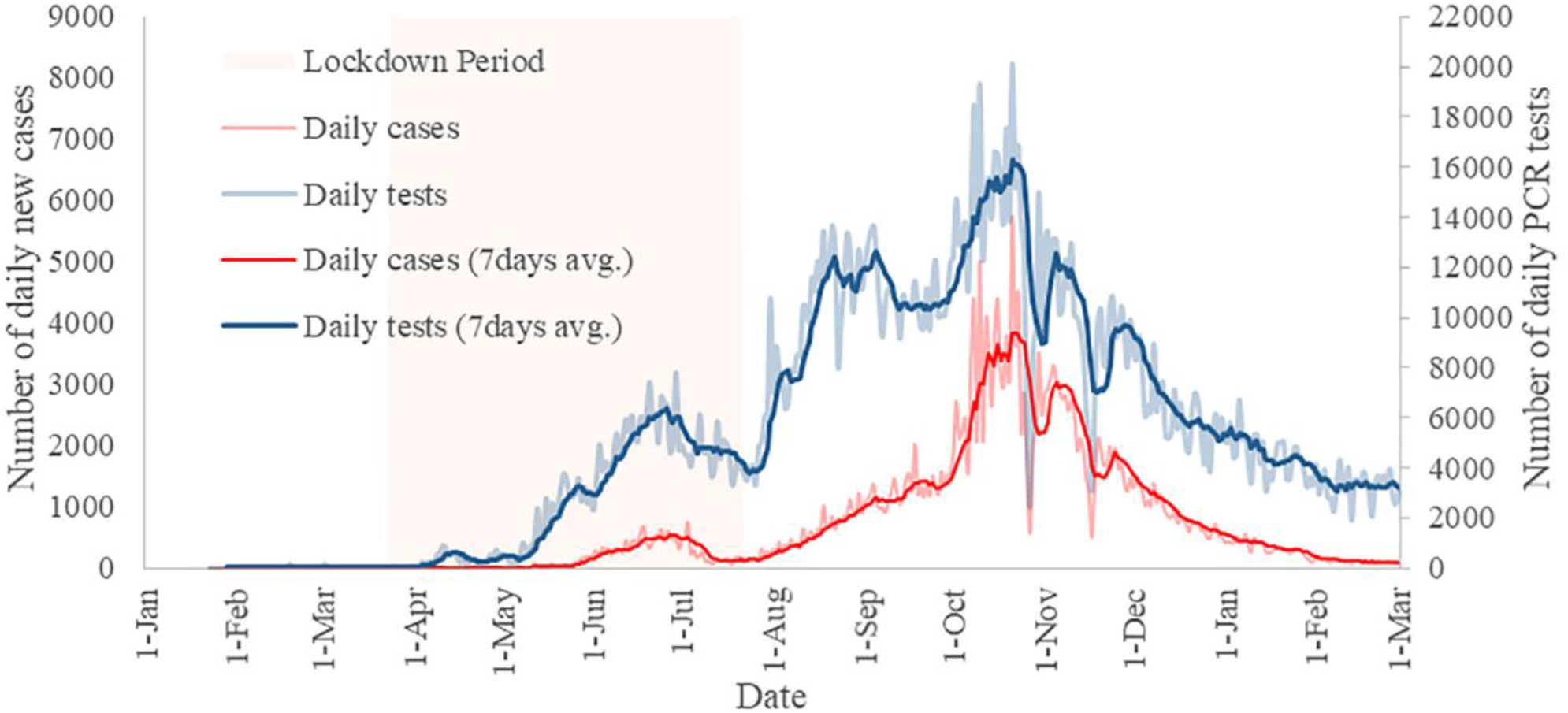
Daily new cases and RT-PCR tests along with 7 days average of cases and tests as of 01 March 2021.

**Figure 8:**
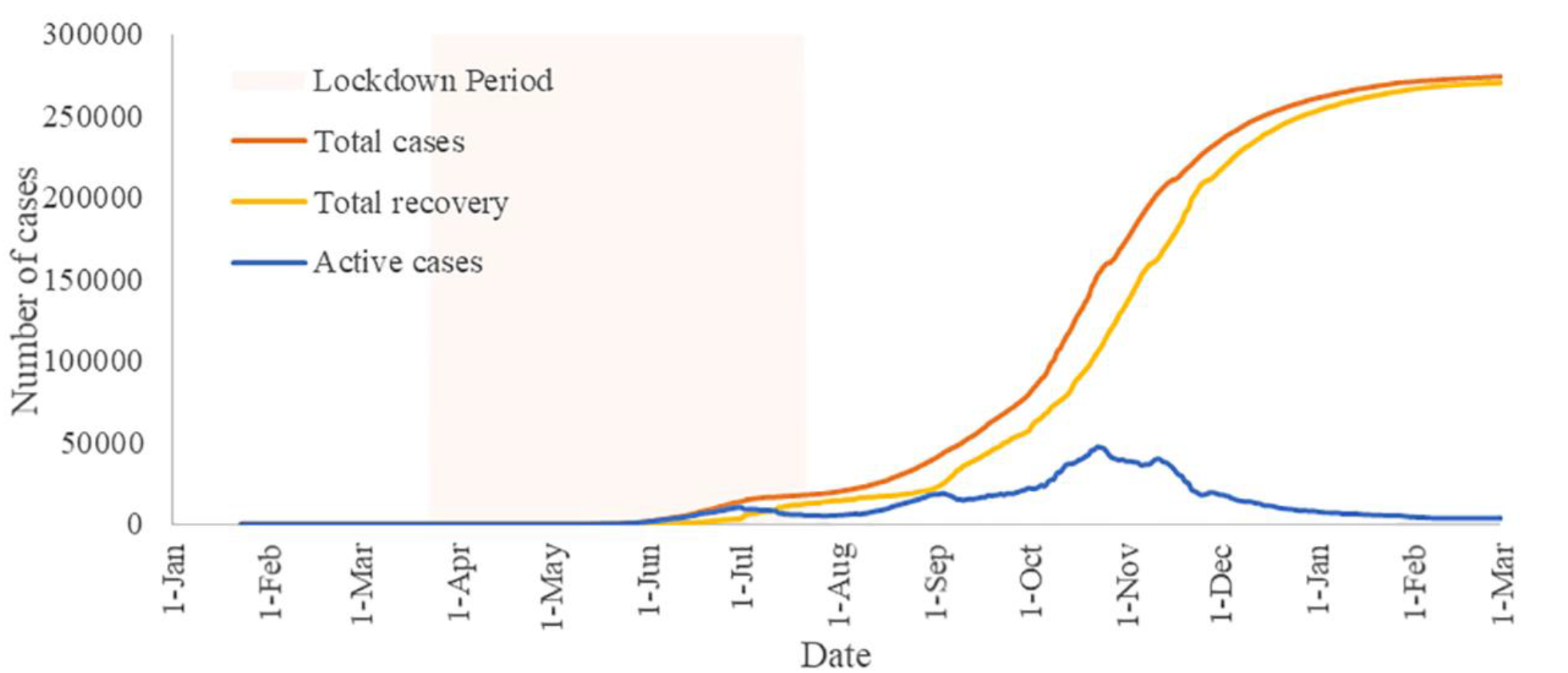
Total cases, recovery, and active cases in Nepal as of 01 March 2021.

Figure 9 shows the distribution of COVID-19 cases among the age group and gender as of 01 March 2021. It shows the community transmission of the coronavirus in the post-lockdown period contributed to the increase of cases among higher age groups and in the female. As discussed previously, one of the challenges is having a higher mortality rate when the virus spread among the older generation, which has been the case in recent days. As of 01 March 2021, mortality rate was above 1%.

**Figure 9:**
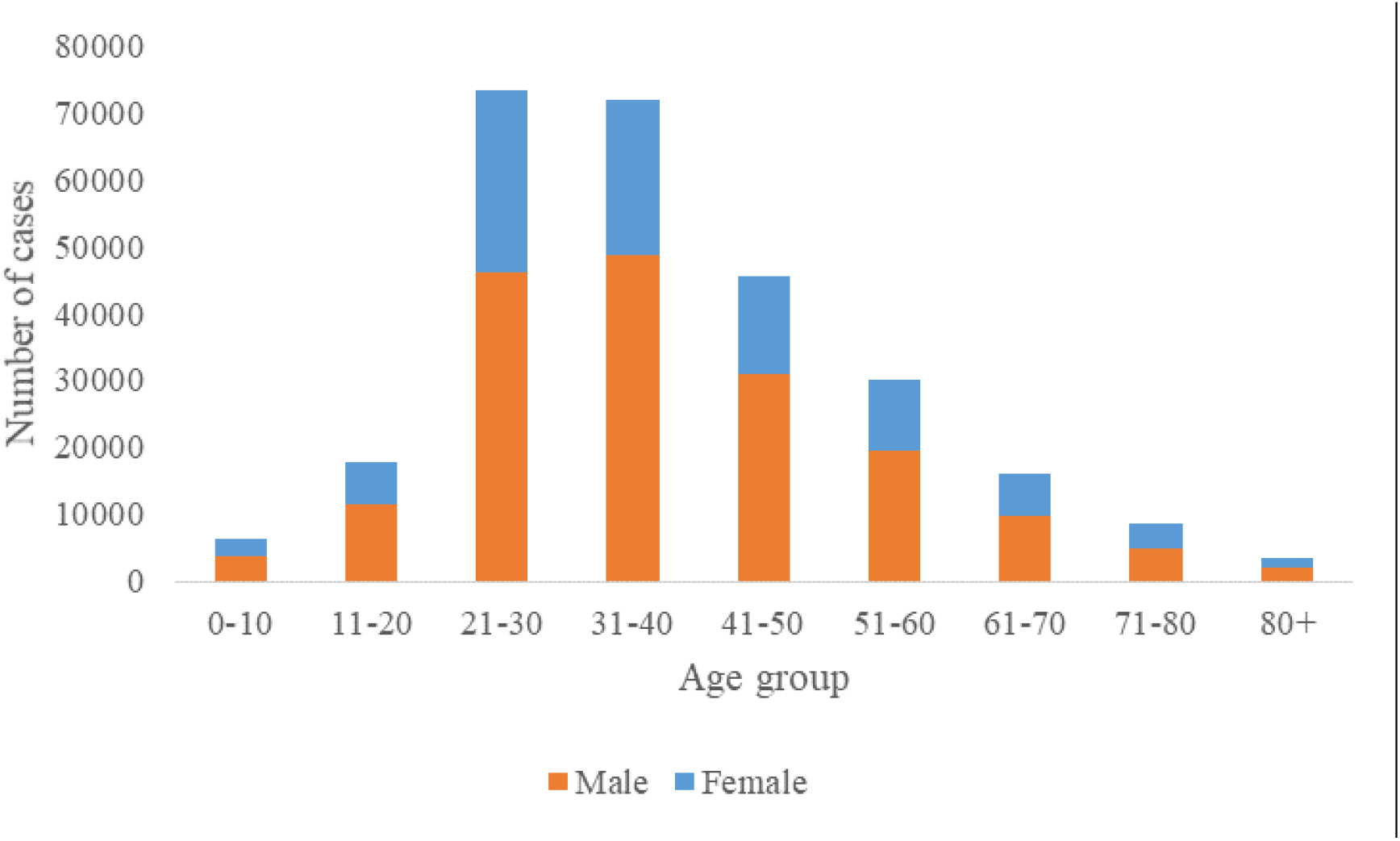
COVID-19 cases distribution among age group and gender as of 01 March 2021.

Figure 10 (A) shows the number of cases in the districts. Compared with the lockdown period, major cities including the capital city Kathmandu have a relatively higher number of cases, which resembles the overall risk scenario reported (15). Figure 10 (B) shows the infected population percentage at the district level across the country. Mugu had a lowest (0.05%) infected population whereas Kathmandu had a maximum of 4.5% infected population.

**Figure 10 (A):**
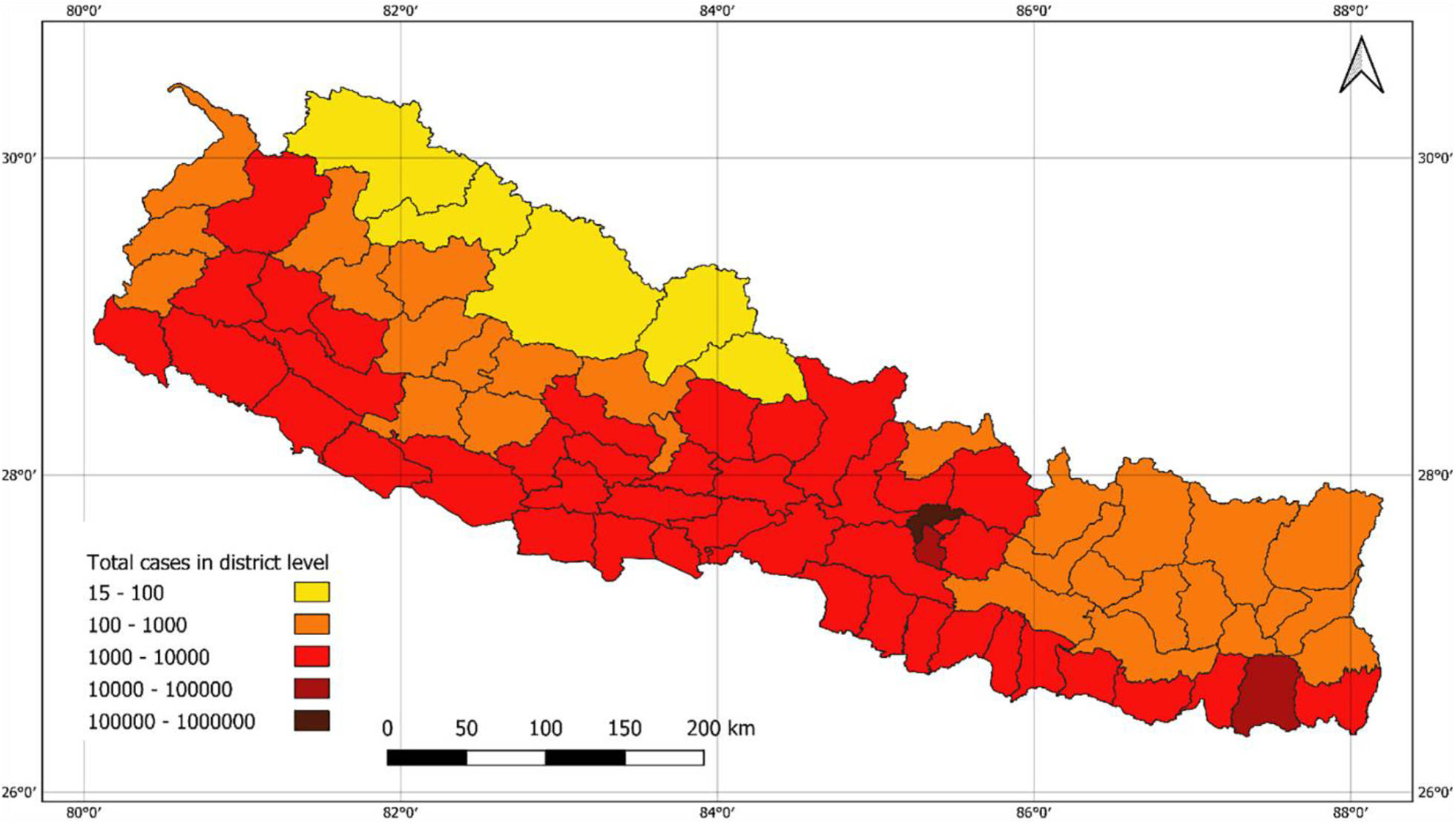
Number of cases in district level as of 01 March 2021.

**Figure 10 (B):**
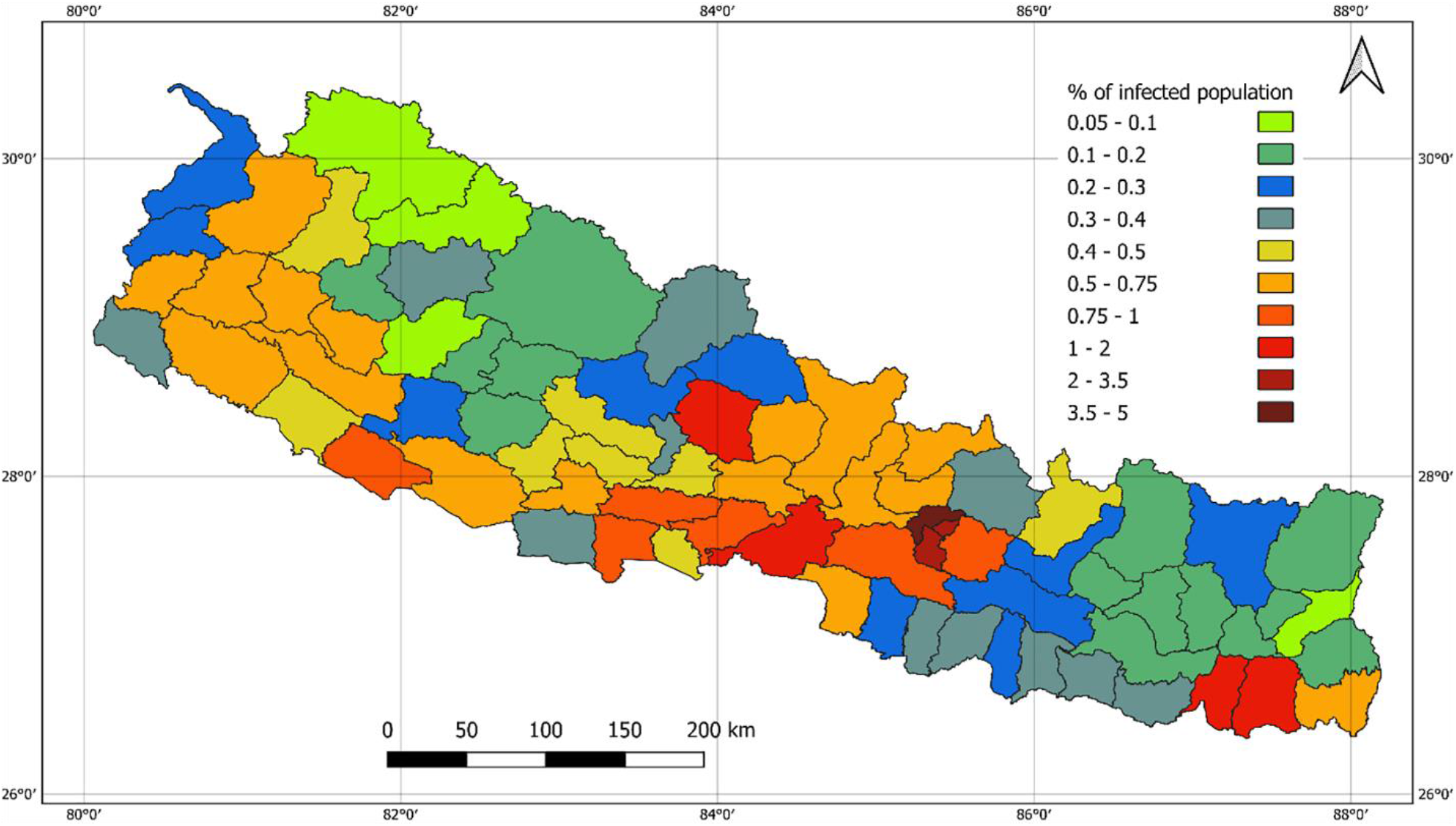
Population infection rate in district level as of 01 March 2021.

## 5 Conclusion and Recommendation

This study provides an overall scenario of the COVID-19 pandemic during the lockdown in Nepal. The capacity of the health system to quickly test to find out if anyone develops symptoms of COVID-19 and tracing and testing of close contacts of those who test positive for COVID-19 must be increased. The government needs to allocate resources, such as the necessary public health workforce, availability of personal protective equipment, expansion of intensive care unit beds, and purchase of extra ventilators. Other actions to stop spread include managed isolation in a designated setting for people who cannot afford to self-isolate or in a dedicated quarantine facility who can’t self-quarantine.

Another approach the government could take to manage local COVID-19 outbreaks is to impose local restrictions. This could be in the form of local lockdown based on risk assessment rather than the nationwide lockdown. As the restrictions of COVID-19 lockdown are eased, there would be more flow of people which leads to more exposure so preventive measures should be established in shopping centres, cities, shops and workplaces. Citizen and institutional/governance awareness is always a key factor in disaster risk reduction and managing pandemic, that often lacking in the developing world (46). The government should continue disseminating information on following social distancing, hand hygiene, and face covering. Data from the post lockdown period also shows the ways to tackle future pandemics considering the socioeconomic condition of the community.

## Data Availability

The authors confirm that the data supporting the findings of this study are available within the article.

## 7 Author Contributions

KS, AB and RRP conceptualised the study. KS and RRP produced most of the figures, analysed, and interpreted the data. KS, AB and RRP prepared the first draft of the paper. All authors have approved the final version.

## 9 Acknowledgements

We acknowledge the Ministry of Health and Population, Nepal for making COVID-19 data publicly available.

## 11 Supplementary Material

None.

## 12 Data Availability Statement

The original contributions presented in the study are included in the article/supplementary material, further inquiries can be directed to the corresponding author.

